# Engineered CRISPR/Cas12a Enables Rapid SARS-CoV-2 Detection

**DOI:** 10.1101/2020.12.23.20248725

**Authors:** Long T. Nguyen, Santosh R. Rananaware, Brianna L.M. Pizzano, Brandon T. Stone, Piyush K. Jain

## Abstract

The coronavirus disease (COVID-19) caused by SARS-CoV-2 has swept through the globe at an unprecedented rate. CRISPR-based detection technologies such as DETECTR, SHERLOCK, and STOPCovid have emerged as a rapid and affordable platform that can shape the future of diagnostics. Recently, we reported engineered crRNAs for Cas12a, called ENHANCE, that enables enhanced detection of nucleic acids. Here we report development, clinical validation, and advancement of ENHANCE platform for detecting SARS-CoV-2. With an RT-LAMP pre-amplification step, ENHANCE detects samples down to a single copy with 95% accuracy and shows high specificity towards various isolates of SARS-CoV-2 against 31 highly similar and common respiratory pathogens. Utilizing LbCas12a-mediated trans-cleavage activity, ENHANCE works robustly in a wide range of magnesium concentration (3 mM-13 mM), allowing for further assay optimization. Additionally, ENHANCEv2 is developed to further improve the previously reported ENHANCE. ENHANCEv2 employs mutated LbCas12a^D156R^, engineered chimeric DNA-extended crRNA, and a dual reporter for both fluorescence-based reporter assay and lateral flow assay. Both ENHANCE and ENHANCEv2 are validated in 62 clinical nasopharyngeal swabs, showing 60/62 (96.7%) agreement with RT-qPCR results, and using only 5 μL of sample and 20 minutes of CRISPR reaction. Lateral flow assay on paper strips displays 100% agreement with fluorescence-based reporter assay in the clinical validation. Following a 30-minute pre-amplification RT-LAMP step, the lyophilized ENHANCEv2 is shown to achieve high sensitivity and specificity while reducing CRISPR reaction time to as low as 3 minutes and maintaining its detection capability upon storage at room temperature for several weeks.

## INTRODUCTION

With a global pandemic of over 72 million COVID-19 cases resulting in over 1.6 million deaths, there remains a crucial need for diagnostic tools that allow for quick yet accurate detection of SARS-CoV-2 without the requirement of expensive equipment and extensive training (1, 2). While vaccines are beginning to emerge from Phase III clinical trials and be granted Emergency Use Authorizations (EUAs) by the FDA (3-7), case numbers are continuing to rise as second and third waves occur in many countries across the globe, and with treatments still somewhat limited, improvements in testing are more necessary than ever to keep both case numbers and fatality numbers down in order for preventative measures to be effective down the line (8, 9).

SARS-CoV-2 is a ∼30kb betacoronavirus composed of four known structural proteins, including the spike (S), nucleocapsid (N), membrane (M), and envelope (E) proteins, as well as a viral RNA genome (10). Nucleic acid detection methods often target N, E, and RdRp (RNA-dependent RNA polymerase) genes. In quantitative Reverse Transcription Polymerase Chain Reaction (RT-qPCR), the viral RNA is reverse transcribed into complementary DNA (cDNA) and then amplified through cyclic changing of temperatures to achieve denaturation of strands, annealing of primers to the template DNA, and extension of new complementary DNA by the addition of nucleotides by a polymerase. DNA copies are detected in real-time through the emission of fluorescence by probes that bind to the DNA. Detection using this method requires expensive equipment, clinically trained professionals and is not easily portable (11, 12).

CRISPR-Cas-based methods, such as SHERLOCK (Specific High sensitivity Enzymatic Reporter unLOCKing) and DETECTR (DNA Endonuclease-Targeted CRISPR Trans Reporter) have recently received an EUA by the FDA. They take advantage of the collateral cleavage (trans) activity from Class 2 Type V and Type VI Cas proteins, specifically Cas13a and Cas12a, to cleave a FRET-based reporter resulting in fluorescence. A Cas12a-based DETECTR has a limit of detection of around 20 copies/μL, while SHERLOCK is based on Cas13a and is time-consuming (∼1 hour, as opposed to ∼30 min) but can detect ∼ 6.75 copies/μL (table S1) (13-20). To amplify the target genes while accommodating for the drawbacks of traditional PCR, both COVID-19 detection platforms pair CRISPR/Cas reactions with an antecedent isothermal amplification step, with SHERLOCK utilizing Recombinase Polymerase Amplification (RPA) and DETECTR utilizing Reverse Transcription Loop Mediated Isothermal Amplification (RT-LAMP). The RT-LAMP, performed at a constant temperature of 60-65°C, is significantly less time-consuming due to its “cauliflower-like” logistic growth amplification pattern. It is also more sensitive than RPA and can be easily adopted without supply-chain issues (21).

Here we report the clinical validation of ENHANCE system, which was developed in our previous study (22), utilizes a similar system with a 7-nucleotide DNA extension on the 3’ end of the crRNA to boost the collateral cleavage activity of an LbCas12a protein, enabling a significant increase in sensitivity while maintaining specificity even at low magnesium concentrations. Similar to DETECTR, RT-LAMP is used for a pre-amplification step prior to the CRISPR-Cas reaction, which trans-cleaves the fluorophore from the quencher after the initial cis-cleavage of the target dsDNA. In order to validate the ENHANCE v1 system under clinical conditions, 62 nasopharyngeal samples (31 positive samples and 31 negative samples) were tested using both the fluorescence-based and lateral flow assays and then compared to RT-qPCR results. Further, we developed a lyophilized a version of ENHANCEv2 system, which utilizes modified crRNAs from ENHANCE, a mutated LbCas12a protein (Cas12a^D156R^) (23-26) to further amplify the signal, and a dual reporter construct that allows each sample to be read using a fluorescence-based and a lateral flow assay format in the same reaction. The lyophilized CRISPR reaction in ENHANCEv2 occurs at an accelerated rate and preserves for a long period of time upon room temperature storage.

## RESULTS

### CRISPR-ENHANCE demonstrates robust detection across SARS-CoV-2 genes

Our previous study observed that a 3’-end chimeric DNA-extended crRNA enhanced the rate of trans-cleavage activity (Kcat/Km) of LbCas12a by 3.2-fold thus facilitated higher sensitivity in nucleic acid detection (fig. 1A), also known as ENHANCE (Enhanced Analysis of Nucleic acids with CrRNA Extensions). This universal 7-mer DNA extension is spacer-independent and has been tested in various nucleic acid targets such as GFP, HCV, HIV, and SARS-CoV-2 (22). To detect SARS-CoV-2 at low copy number, both wild-type and ENHANCE require a pre-amplification step such as Reverse Transcription Loop-mediated Isothermal Amplification (RT-LAMP), which reverse transcribes SARS-CoV-2 genomic RNA to complementary cDNA and convert it into dsDNA targets (14, 18).

**Fig. 1.**
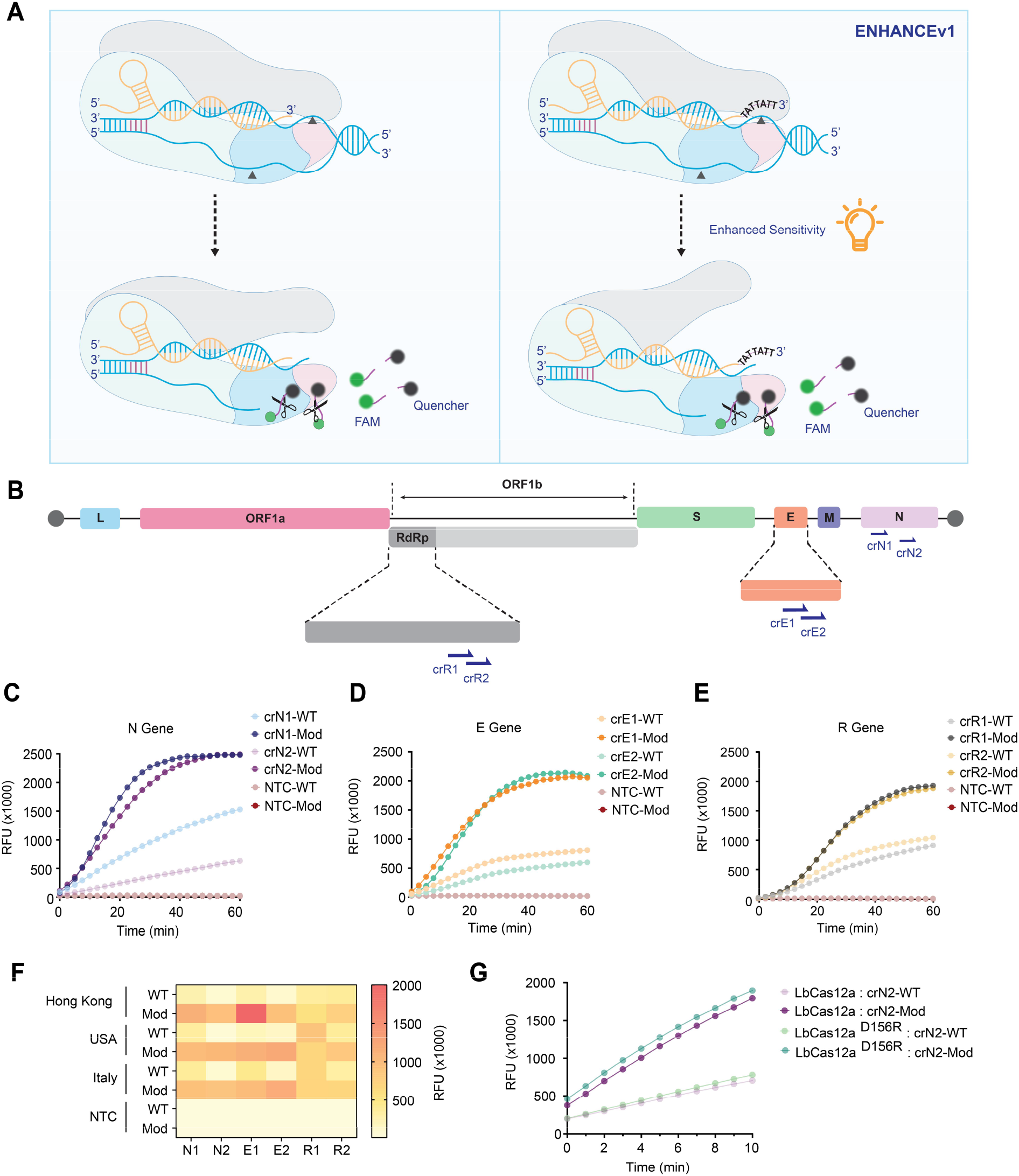
crRNA screening and optimization of ENHANCE for detection of SARS-CoV-2. (**A**) Schematic of detection platform using CRISPR-Cas12a highlighting the differences between wild-type and ENHANCE systems. (**B**) Schematic of SARS-CoV-2 RNA genome and target regions of crRNA used for ENHANCE within the SARS-CoV-2 genomic RNA. (**C**), (**D**), and (**E**) Average fluorescence intensity in RFU of wild-type crRNA (crRNA-WT) and modified crRNA (crRNA-Mod) observed by fluorescence-based reporter assay for N gene, E gene, and RdRp gene, respectively (n = 6). (**F**) SARS-CoV-2 Genomic RNAs obtained from different geographic regions were tested with CRISPR-Cas12a systems. (**G**) Comparison of fluorescence intensities when the mutated LbCas12a^D156R^ is introduced (n = 6). All targeted sequences are genomic RNA obtained from BEI Resources. All fluorescence intensities shown were taken at t = 20 minutes.

In this study, selected regions of the SARS-CoV-2 N gene, E gene, and RdRp gene were targeted with pairs of crRNAs (crN, crE, and crR referred to as crRNA targeting N gene, E gene, and R gene, respectively). These designed crRNAs bear a spacer region of greater than 50% GC content (fig. 1B and table S2). In addition to previously designed crN2, crE1, and crE2 by Broughton et al. (14), we employed three additional crRNAs (crN1, crR1, and crR2) to not only determine optimal crRNAs but also examine the modified LbCas12a trans-cleavage activity compared to the wild-type system. Consistent with our previous study, when targeting SARS-CoV-2 genomic RNA, the modified crRNAs (referred to as crRNA-Mod) exhibited enhanced trans-cleavage activity, up to 5-fold, across all targets compared to the wild-type crRNA (crRNA-WT), indicating significantly higher fluorescence signals via the fluorophore-quencher-based reporter assay results (figs. 1C-E). Improved activity and specificity remained consistent while detecting multiple geographically distributed isolates (Hong Kong, USA, and Italy) (fig. 1F). The ENHANCE system works robustly with a wide range magnesium concentration (3 mM-13 mM), and especially at low magnesium concentration. Thus, ENHANCE enables a potential strategy for screening a library of guide RNAs under various reaction conditions (27).

Previous study has shown that the variant LbCas12a^D156R^ exhibited a substantially higher gene editing efficiency compared to the wild-type LbCas12a (23-26). We hypothesized that this mutated LbCas12a^D156R^ would likely increase the trans-cleavage activity due to its correlation between cis-cleavage and trans-cleavage capabilities. Having previously modified crRNA for enhanced LbCas12a trans-cleavage activity, we sought to investigate if we could utilize LbCas12a^D156R^ to further increase the sensitivity for nucleic acid detection purposes. As a result, the combination of LbCas12a^D156R^ and 7-mer DNA-extended crRNA showed a significantly higher fluorescence sensitivity than the wild-type LbCas12a and crRNA and a slight enhancement compared to the combination of wild-type LbCas12a and modified crRNA (fig. 1G). This observation later became the basis for the development of ENHANCEv2.

### CRISPR-ENHANCE SARS-CoV-2 targeted guide RNA optimization

To seek optimal crRNAs for the ENHANCE platform, we next evaluated the limit of detection (LoD) for the 6 selected modified crRNAs. Quantitative qPCR SARS-CoV-2 genomic RNA were utilized to make a serial dilution theoretically ranging from 200 copies/μL to 0.2 copies/μL followed by an isothermal pre-amplification RT-LAMP step (1000 copies/rxn to 1 copy/rxn). An estimated LoD was determined based on a criterion that the lowest copies/μL having 2/3 of replicates showing at least 5 folds in fluorescence signal compared to that of the no-template control (NTC) within 20 minutes. Out of the six modified crRNAs, crN2 and crE2 designed by Broughton et. al. exhibited the lowest LoD with 0.2 copies/μL and 7.9 copies/μL, respectively (figs. 2A-D). Hence, we selected crN2-Mod and crE2-Mod as the optimal guide RNAs for our clinical validation. The LoDs of crN2-Mod and cRE2-Mod were confirmed by testing 20 replicates of their 1x concentration (0.2 copies/μL and 7.9 copies/μL for crN2 and crE2, respectively) and 1x concentration (0.4 copies/μL and 15.8 copies/μL for crN2 and crE2, respectively). The LoD was established by observing 95% (19/20 replicates) of the target samples being positive. The replicates of both crN2-Mod and crE2-Mod at 1x concentration yielded sufficient positive results, reconfirming the LoD to be 0.2 copies/μL and 7.9 copies/μL, respectively, within 20 minutes of CRISPR reaction (fig. 2E and F). These results demonstrate that the CRISPR-ENHANCE achieved a comparable sensitivity to traditional RT-qPCR testing of SARS-CoV-2 in less than an hour without sophisticated equipment.

**Fig. 2.**
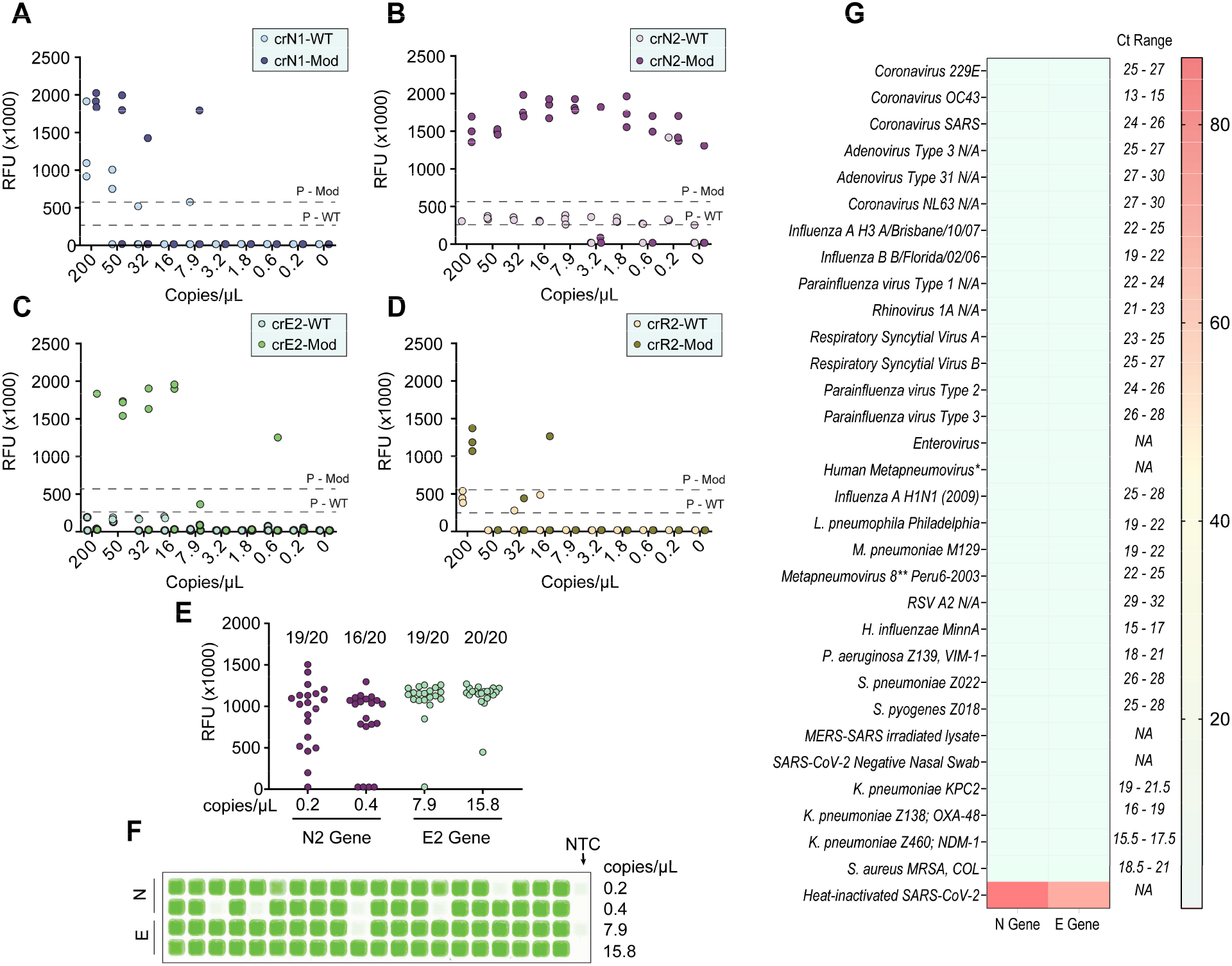
Limit of Detection and Exclusivity Testing of ENHANCE. (**A**), (**B**), (**C**), and (**D**) Fluorescence intensities in RFU of wild-type crRNA (crRNA-WT) and modified crRNA (crRNA-Mod) in serially diluted samples containing quantitative genomic RNA of SARS-CoV-2 (n = 3). (**E**) 2/3 of samples detected in (A), (B), (C), and (D) were subject to be tested with 20 different serial dilutions of 1x and 2x number of copies/μL. (**F**) Scanned images of samples in (E). (**G**) Exclusivity testing of ENHANCE against highly similar and commonly circulating pathogens. Fluorescence signals were taken at t = 20 minutes. All samples with Ct value ranges were obtained from Zeptometrix.

As a part of exclusivity, we tested a variety of highly similar pathogens to SARS-CoV-2 (SARS-CoV, MERS-CoV, Coronavirus 229E, OC43, SARS, and NL63) and respiratory pathogens commonly detected in nasal swabs or saliva. Notably, the CRISPR-ENHANCE exhibited high specificity towards SARS-CoV-2 with the modified crRNAs targeting N gene and E gene (fig. 2G). No cross reactivity was observed in any of the 31 tested pathogens.

### Validation of ENHANCEv1 in patient samples

Having tested detection using the SARS-CoV-2 heat-inactivated virus and genomic RNA for crRNA optimization and LoD determination, we sought to validate the ENHANCEv1 in patient samples to determine whether it still maintains robust detection with clinical conditions. We tested a total of 62 nasal swab samples in which 31 samples were predetermined SARS-CoV-2 positives and the other 31 samples negative. Samples were randomly selected out of the pool and blinded prior to viral RNA extraction. Similar to DETECTR (14), RNAse P gene was used as an internal control for all the nasal swab samples. For the real-time fluorescence-based reporter detection, the criterion for a positive sample is a change in fluorescence signal of 5-fold or greater compared to non-template control (NTC) samples within 20 minutes. Consistent with previous observation, the EHANCEv1 demonstrated a higher sensitivity across all positive samples at low magnesium concentration in a shorter amount of time (fig. 3A). Furthermore, the ENHANCEv1 achieved a 96.7% overall accuracy with a false positive rate and a false negative rate of 3.3% and 3.3%, respectively (fig. S1 and table S3). For lateral flow-based detection, the ENHANCEv1 showed 100% in agreement with the real-time fluorescence-based reporter assay. Similarly, the false positive and false negative rates for this assay were both 3.3% (figs. 3B and S2, S3).

**Fig. 3.**
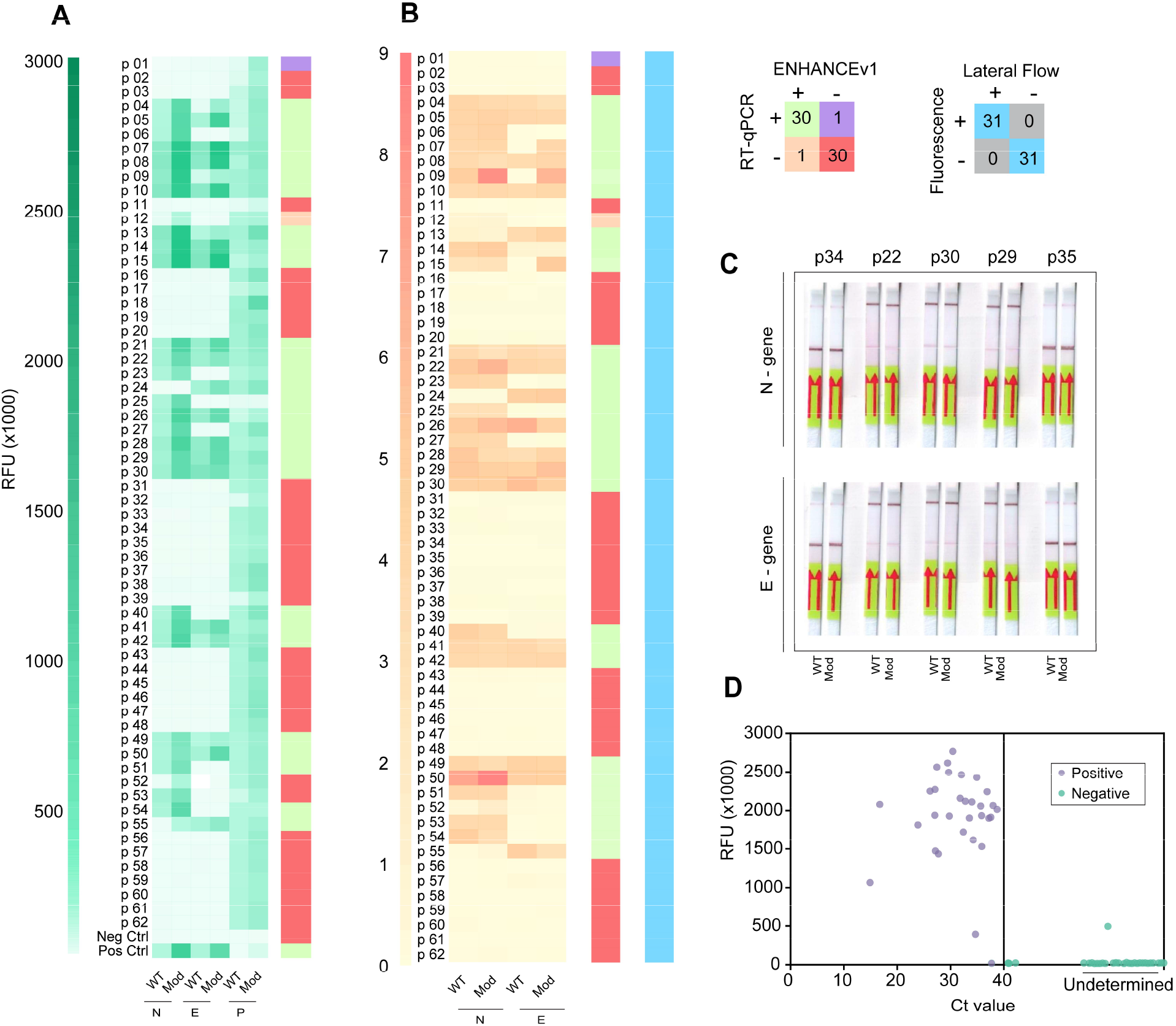
Clinical validation of ENHANCE. (**A**) Fluorescence intensity of wild-type crRNA (crRNA-WT) and modified crRNA (crRNA-Mod) detecting SARS-CoV-2 RNA in 62 patient samples using the fluorescence-based reporter assay. Fluorescence signals were taken at t = 20 minutes. (**B**) Lateral flow assay detecting SARS-CoV-2 RNA in the same 62 patient samples. Heat map shows ratio of band intensity of positive line (top band) to control line (bottom band). Band intensities were analyzed by ImageJ. (**C**) Representation of lateral flow assay testing 62 patient samples. For full detail, please see supplementary materials. (**D**) Maximum fluorescence intensity of 62 patient samples against Ct values validated with RT-qPCR.

### Lyophilized ENHANCEv2 significantly reduces CRISPR reaction time

We further developed a CRISPR-ENHANCEv2 by combining ENHANCEv1 with the mutated LbCas12a^D156R^ and a dual reporter comprising a fluorophore FAM, biotin, and a quencher. This reporter can be used as a fluorescence-based reporter as well as a lateral flow-based reporter, both in a single detection assay (fig. 4A). A typical CRISPR reaction requires complexation of crRNA and LbCas12a prior to addition of pre-amplified activator. Therefore, we initially sought to understand whether the complexation of crRNA and LbCas12a can be achieved and remain stable for a period of time prior to testing. If successful, large batches of pre-complexed crRNA:LbCas12a can be manufactured to reduce reaction time and minimize the need for specialized training. We explored if lyophilization can accomplish this objective. Optimal concentrations of crRNA and LbCas12a^D156R^ were pre-complexed in a mixture containing dual reporter and cleavage buffer. This combined mixture was prepared in different pilot scales: 1 reaction, 5 reactions, and 20 reactions (fig. 4B). By conducting a time study, we observed that these lyophilized pre-complexes maintain high trans-cleavage activity as the liquid version and remain stable for up to 30 days upon storage at room temperature (figs. 4C and S5A). These results help enable scale-up manufacturing, ease of transportation, storage, and reduction in preparation time.

**Fig. 4.**
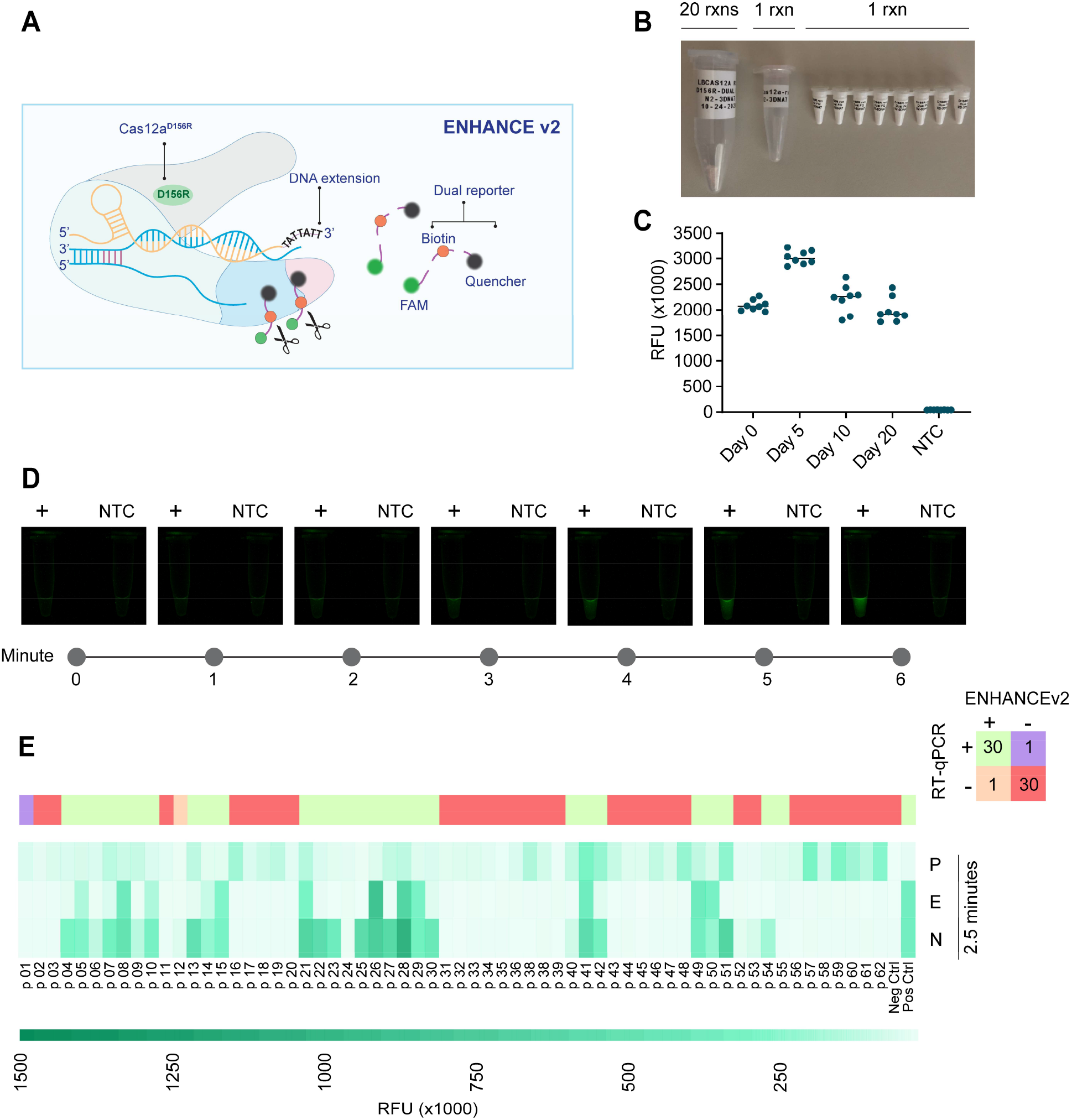
Clinical validation of ENHANCEv2. (**A**) Schematic diagram of composition of ENHANCEv2 system. (**B**) Lyophilized samples in different pilot scales (20, 5, and 1 reactions) of CRISPR reaction used in ENHANCEv2. The CRISPR reaction contains pre-complexed crRNA:LbCas12a^D156R^, dual reporter and NEBuffer 2.1. Only RNase-free water and RT-LAMP products are added to activate the reaction. (**C**) Stability testing of lyophilized CRISPR reaction using ENHANCEv2 with respect to time. (**D**) Time-course observation of lyophilized CRISPR reaction. Positive samples can be seen by naked eyes under blue light within as low as 2 minutes. (**E**) Fluorescence intensity of ENHANCEv2 detecting 62 patient samples at t = 2.5 minutes.

Surprisingly, we also observed a significant decrease in reaction time. Using lyophilized CRISPR reaction stored at −20°C after 2.5 months for the fluorescence reporter assay, the fluorescence signal can be seen by naked eyes under blue light after 2 minutes (fig. 4D). When applying the lyophilized ENHANCEv2 towards 62 patient samples, we were able to accurately detect the same 62 patient nasal swab samples with >5-fold change in fluorescence compared to NTC samples in as low as 2.5 minutes (fig. 3F and S4), bringing a total reaction time to 33 minutes (30 minutes of RT-LAMP). This phenomenon strongly suggests that pre-complexed LbCas12a-crRNA promotes faster and easier assay setup.

## DISCUSSION

In the wake of the COVID-19 outbreak, detection and vaccine developments become the two important pillars of research to eradicate the disease (28, 29). At the time of preparing this manuscript, the first vaccine developed by Pfizer has been granted emergency authorization by the U.S. Food and Drug Administration. This news gives hope to combat the COVID-19 disease that has already caused great damage to our community. However, it will take months for the vaccine to get in the hands of million people; therefore, COVID-19 testing remains a necessary tool to track the severity of the spread and to follow up the population post-vaccination. Many CRISPR-based detection platforms such as DETECTR, SHERLOCK, and STOPCovid have emerged as an alternative to traditional RT-qPCR assays due to their rapid, affordable, and sensitive capabilities (14, 18, 30). Here, we developed ENHANCEv2, an engineered CRISPR-based diagnostic platform to detect SARS-CoV-2 in patient samples with high sensitivity and specificity. Similar to the DETECTR system, ENHANCE utilized a robust trans-cleavage capability of Cas12a towards detection (13, 14).

ENHANCEv1 consists of a chimeric 7-mer TA-rich extension the 3’-end of crRNA and wild-type LbCas12a. Our previous study has shown that this modified CRISPR-Cas system significantly enhances Lbcas12a trans-cleavage activity while slightly improving its specificity (22). The theoretical limit of detection for ENHANCEv1 was determined to be 0.2 copies/μL and 7.9 copies/μL for N gene and E gene, respectively, achieving single molecule sensitivity in a sample volume as small as 5 μL with 95% accuracy. This engineered system also exhibited high specificity towards SARS-CoV-2 when tested against highly similar and commonly circulating pathogens. Finally, ENHANCEv1 was used to validate 62 patient samples in this work, with 60/62 patients in agreement with RT-qPCR results. One false negative and one false positive were observed in 31 negative and 31 positive samples for both fluorescence-based reporter and lateral flow assay, showing a total of 96.7% accuracy. We encountered some challenging results during clinical validation of ENHANCEv1 and ENHANCEv2 with 62 patient samples. Patient sample p01 was pre-determined as negative upon arrival in our lab. The sample also indicated as negative with our ENHANCE assay, but revalidation with RT-qPCR assay indicated it as a positive sample. Therefore, we concluded that the sample was a false negative based on our analysis approach. For patient sample p43, both ENHANCE and RT-qPCR assays agreed that the sample was negative with SARS-CoV-2, but it was pre-determined as a positive sample. Thus, we concluded this sample was negative. The patient sample p12 was predetermined to be a negative sample and was detected negative with the qPCR, however, it was consistently detected as positive by ENHANCE v1 and v2 methods. Therefore, we concluded it as a false positive. Finally, QC failure was observed while testing samples p32, p38, and p62 as fluorescence signals for the RNase P control gene did not show up. However, we were able to successfully detect these samples upon the second round of RNA extraction.

We developed ENHANCEv2 to further improve the testing capability of ENHANCEv1. ENHANCEv2 is comprised of a 7-mer DNA extension on the 3’-end of crRNA, a mutated LbCas12a^D156R^, and a dual reporter that can be used for both fluorescence-based reporter and lateral flow assays. The dual reporter enables detection of clinical samples in two different assay formats that can be potentially used for conducting a high-throughput lab-based as well as a rapid point-of-care-based test using a single CRISPR kit (fig. S5B). ENHANCEv2 was used to test the same 62 patient samples, showing 100% agreement with ENHANCEv1 while reducing CRISPR reaction time to as low as 3 minutes. Furthermore, unlike ENHANCEv1, the ENHANCEv2 does not require a crRNA/Cas pre-incubation step, saving 15-20 minutes of time and labor. These results were achieved by lyophilizing pre-complexed crRNA:LbCas12a in advance. The lyophilized version of ENHANCEv2 not only accelerates CRISPR reaction speed significantly but also remains stable for several weeks upon storage at room temperature.

Similar to SHERLOCK and DETECTR, a major drawback to this technology is the possibility of carryover contamination. CRISPR-based detection system provides two check points for detection: (1) an isothermal pre-amplification step such as RT-LAMP that converts SARS-CoV-2 genomic RNA into millions of dsDNA copies and (2) CRISPR reaction where crRNA:LbCas12a complex targets the amplified dsDNA. Since this detection platform is not a continuous step, contamination might occur. Therefore, proper reaction setup must be met. This major disadvantage motivated us to develop ENHANCEv2. ENHANCEv2 uses RT-LAMP master mix containing UDG which has been shown to greatly eliminate carryover contamination. The lyophilized CRISPR version not only significantly reduces the reaction time but also simplifies the needs for CRISPR reaction setup. The reaction can be reconstituted with the addition of RNase-free water and RT-LAMP products. The method further promotes large-scale manufacturing, eases the challenge of reagent storage and handling, and would enable easier deployment across the globe.

## CONCLUSIONS

The ENHANCEv2 offers a rapid, sensitive, and affordable test for detecting SARS-CoV-2 in clinical samples. The improved capability and stability of ENHANCEv2 would be pivotal for global deployment towards curbing the COVID-19 pandemic.

## MATERIALS AND METHODS

### Protein Expression and Purification

LbCas12a gene fragment was obtained by PCR from the plasmid LbCpf1-2NLS as a gift from Jennifer Doudna (Addgene plasmid # 102566) (31) and subcloned into a linearized CL7-tagged vector (Plasmid #21, TriAltus Bioscience) digested with HindIII and XhoI. The fragments assembly was performed using NEBHiFi Builder (New England Biolabs) following the manufacturer’s protocol. The assembled plasmid was transformed into DH5α (New England Biolabs) competent cells. Individual colonies were picked the next day and inoculated in 10 ml of LB broth, Miller (Fisher Scientific) at 37°C overnight. Cells were harvested, and plasmids were extracted and purified using the Monarch Mini Plasmid prep (New England Biolabs).

For protein expression, 100 ng of the purified plasmid was transformed into BL21(DE3) competent cells (New England Biolabs). Individual colonies were picked and inoculated in 10 ml of Terrific Broth (TB) at 37^°^C for 8-10 hours. The culture was then added to 1L TB broth containing 50 μg/ml Kanamycin (Fisher Scientific) and 50 μL antifoam 204 (Sigma Aldrich) and let grown until OD600 = 0.6. The culture was then taken out of the 37^°^C incubator and let cooled on ice for 30-45 minutes. Isopropyl β-D-1-thiogalactopyranoside (IPTG, Fisher Scientific) was added to the culture and let grown at 18^°^C overnight.

The overnight culture was centrifuged to collect cell pellets the next day. They were resuspended in Lysis Buffer A (2M NaCl, 50 mM Tris-HCl, pH = 7.5, 0.5 mM TCEP, 5% Glycerol, 1mM PMSF, 0.25 mg/ml lysozyme). The mixture was disrupted by sonication, centrifuged at 40000 x g, and filtered through a 0.45 μm filter. The lysate was run through a 1 ml CL7/Im7 column (TriAltus Bioscience) connected to the FPLC Biologic Duoflow system (Bio-Rad). The column was washed for at least 3 cycles of alternating Wash Buffer B (2M NaCl, 50 mM Tris-HCl, pH = 7.5, 0.5 mM TCEP, 5% Glycerol) and Wash Buffer C (50 mM Tris-HCl, pH = 7.5, 0.5 mM TCEP, 5% Glycerol). The column was then eluted by adding 5 mL of SUMO protease (purified from plasmid pCDB302 as a gift from Christopher Bahl, Addgene plasmid# 113673) (32) and flown through in a closed-loop cycle at 30^°^C for 1.5 hours. (Optional) To remove SUMO protease, the eluted solution was then concentrated using a 30 kDa MWCO Sartorius Vivaspin Concentrator to 500 μL and subject to size exclusion chromatography in SEC buffer (500 mM NaCl, 50 mM Tris-HCl, pH = 7.5, 0.5 mM TCEP) via the Superdex 200 increase 10/300 GL column (Cytiva). Eluted fractions were collected, pooled together, concentrated, quantified using the NanoDrop (Thermo Fisher), snap frozen in dry ice, and stored at −80^°^C until use.

For LbCas12a^D156R^ expression and purification, the CL7-tagged plasmid obtained by subcloning above was mutated using the Q5® Site-Directed Mutagenesis Kit (New England Biolabs) following the manufacturer’s protocol. The protein expression and purification were the same as described above.

### Lyophilization of ENHANCEv2 CRISPR reaction

To make one CRISPR reaction, reagents were mixed in the following order:

| Reagent | Volume | Final concentration |
| --- | --- | --- |
| 100 μM LbCas12a<sup>D156R</sup> | 0.05 μL | 100 nM |
| 100 μM crRNA-Mod | 0.0625 μL | 125 nM |
| 100 μM Dual reporter | 0.25 μL | 500 nM |
| 10x NEBuffer 2.1 | 5 μL | 1X |
| RNase-free water | 44.6375 μL | |
| **Total** | **50 μL** |  |

This recipe was scaled up accordingly to make 5x and 20x reaction aliquots. These aliquots were then subject to lyophilization using the Labconco freeze dryer for 2-4 days.

### CDC RT-qPCR assay

Patient samples were obtained from University of Florida (UF) Clinical and Translational Science Institute and Boca Biolistics by following the guidelines listed in the approved UF Institutional Review Board protocol (IRB202000781). The samples were re-validated with real time RT-qPCR. The reactions were performed using the CDC recommended Quantabio qScript XLT One-Step RT-qPCR ToughMix (Catalog# 95132-100) and TaqMan probes and primer sets (add citation) and measured using the ViiA 7 Real-Time PCR System. RT-qPCR quantification was performed using amplification plots generated by the ViiA 7 software.

### Viral nucleic acid extraction

For crRNA screening and optimization, LoD estimation, inclusivity testing and specificity testing, viral RNA extraction was performed using the Lucigen QuickExtract™ DNA Extraction Solution (Cat # QE09050). Viral samples were diluted with QuickExtract™ in a 1:1 (v/v) ratio and incubated at 65°C for 15 min and 98°C for 2 min.

For clinical validation experiments, all patient samples were extracted using Maxwell® RSC 16 automated nucleic acid extraction instrument. Maxwell® RSC Viral Total Nucleic Acid Purification Kit (Cat# AS1330, as recommended by CDC) was used for all extractions following manufacturer’s protocol.

### RT-LAMP reactions

A set of 6 LAMP primers were designed for each gene using the freely available PrimerExplorer software (https://primerexplorer.jp/e/) (33). The designed primers were synthesized by Integrated DNA Technologies. A 10x primer mix for each gene was created by mixing the 6 primers to a final concentration of 1.6 μM (FIP/BIP), 0.8 μM (LB/LF) and 0.2 μM (F3/B3). RT-LAMP reactions were performed using the WarmStart® Colorimetric LAMP 2X Master Mix (New England Biolabs). RT-LAMP master mix for N reactions (including the positive and negative control) were prepared as follows:

|  | 1x reaction | (N+1)x reactions |
| --- | --- | --- |
| WarmStart Colorimetric LAMP 2X Master Mix with UDG | 12.5 μL | (N+1) x 12.5 μL |
| LAMP Primer Mix (10X) | 2.5 μL | (N+1) x 2.5 μL |
| Target RNA | 5 μL | (N+1) x 5 μL |
| Nuclease Free Water | 5 μL | (N+1) x 5 μL |
| **Total** | **20 μL** | **(N+1) x 20 μL** |

The prepared master mix was incubated at 65°C for 30 min to allow for adequate amplification. The amplified products were tested downstream using the CRISPR ENHANCE assays.

### crRNA screening and optimization of ENHANCE for detection of SARS-CoV-2

Genomic RNA from SARS-CoV-2, Isolate USA-WA1/2020 (NR-52285) obtained from Biodefense and Emerging Infections Research Resources Repository (BEI resources), was spiked in nucleic acid extract obtained from a healthy donor. The spiked extract was amplified for the target genes (N1, N2, E1, E2, R1 and R2) using RT-LAMP. The amplified products were then detected using wild type CRISPR/Cas12a as well as CRISPR ENHANCEv1.

### Inclusivity Testing

SARS-CoV-2 Genomic RNA of isolates obtained from different geographic regions such as Italy, Hong Kong and USA (NR-52498, NR-52388 and NR-52285) obtained from BEI resources were spiked in nucleic acid extract obtained from the nasopharyngeal swab of a healthy donor. Target genes were amplified using RT-LAMP protocol described earlier and detected using CRISPR ENHANCEv1.

### Specificity testing

To demonstrate specificity of our assay towards SARS-CoV-2 we obtained 31 highly similar and commonly circulating pathogens from ZeptoMetrix (Cat# NATPPQ-BIO, Cat# NATRVP-3, Cat# NATPPA-BIO). Each pathogen was spiked in a matrix composed of nasopharyngeal swab from a healthy donor. The spiked nasal swab was extracted using Lucigen QuickExtract™ and target genes within the extracted nucleic acids were amplified using RT-LAMP. The amplified products were detected using CRISPR ENHANCEv1.

### Estimation of LoD

Nasopharyngeal swab sample from a healthy donor was mixed with an equal volume of Lucigen QuickExtract™ DNA Extraction Solution (Cat # QE09050) and incubated at 65°C for 15 min and 98°C for 2 min to extract nucleic acids from the swab. Mock clinical patient samples were prepared by serially diluting the nucleic acid extract with Quantitative PCR (qPCR) Control RNA from Heat-Inactivated SARS-Related Coronavirus 2 (BEI NR-52347) to a final concentration range of 200 copies/μL to 0.2 copies/μL. The LoD for each gene was determined by amplifying the gene using RT-LAMP and then detecting it at the indicated concentrations with CRISPR ENHANCEv1. The LoDs for N2 gene and E2 gene were confirmed by testing with 20 replicates at 1x and 2x of the previously estimated LoD for each gene.

### Fluorescence-based reporter detection assay

All fluorescence-based detection experiments were performed in a 384-well plate. The following recipe was used for CRISPR/Cas complexation:

|  | 1x reaction | (N+1)x reactions |
| --- | --- | --- |
| 3 μM LbCas12a | 0.4 μL | (N+1) x 0.4 μL |
| 3 μM crRNA | 0.8 μL | (N+1) x 0.8 μL |
| 10X NEBuffer 2.1 | 1.2 μL | (N+1) x 1.2 μL |
| Nuclease-free Water | 9.6 μL | (N+1) x 9.6 μL |
| **Total** | **12 μL** | **(N+1) x 12 μL** |

The prepared master-mix was incubated at 37°C for 15 min after which it is added to a 384-well plate containing 0.2 μL of 100 μM Fluorophore-Quencher (FQ), 25.8 μL of nuclease-free water and 2 μL of RT-LAMP product. Emitted fluorescence resulting from Cas12a based trans-cleavage was measured using BioTek Synergy 2 microplate reader with fluorescence measurement at excitation and emission wavelengths of 485/20 and 528/20, respectively, every 2.5 minutes. For a 96-well plate format all reagents are scaled up 2.5 times.

### Lateral Flow detection assay

The following recipe is used for the lateral flow assay:

|  | 1x reaction | (N+1)x reactions |
| --- | --- | --- |
| 3 μM LbCas12a | 0.5 μL | (N+1) x 0.5 μL |
| 3 μM crRNA | 1 μL | (N+1) x 1 μL |
| 10X NEBuffer 2.1 | 5 μL | (N+1) x 5 μL |
| 5 μM FAM-Biotin reporter | 2 μL | (N+1) x 2 μL |
| Nuclease-free Water | 39.5 μL | (N+1) x 39.5 μL |
| **Total** | **48 μL** | **(N+1) x 48 μL** |

2 μL of the corresponding RT-LAMP product was added to the above mixture and incubated at 37°C for 20 minutes. A paper strip is then dipped in the reaction tube and the presence or absence of the target gene is determined based on a visual readout after 2 mins. Lateral flow band signals were later quantified by ImageJ.

### Clinical validation of patient samples using CRISPR ENHANCE

62 patient samples underwent a blind test. Random samples were selected from the pool, and nucleic acids extracted from those patient samples were subjected to RT-LAMP based amplification of N2 gene, E2 gene and RNASE-P gene. The same amplified RT-LAMP products were then detected using both the fluorescence-based reporter detection assay and the lateral flow assay with ENHANCEv1 and with the lyophilized ENHANCEv2.

## Data Availability

All data associated with this study are in the main text and the Supplementary Materials. The CL7-tagged LbCas12a and LbCas12aD156R expression plasmid will be soon deposited to Addgene after obtaining the required permissions.

## SUPPLEMENTARY MATERIALS

Fig. S1. Clinical validation of ENHANCE for the detection of SARS-CoV-2 in 62 patient samples using lateral flow assay.

Fig. S2. Quantitative analysis of lateral flow assay using ENHANCE on 62 patient samples.

Fig. S3. Clinical validation of ENHANCEv2 for the detection of SARS-CoV-2 in 62 patient samples using fluorescence-based reporter assay.

Fig. S4. Clinical validation of SARS-CoV-2 detection in 62 patient samples using RT-qPCR. Table S1. Comparison of CRISPR-based detection methods.

Table S2. Sequences used in this study. Table S3. Interpretation of results.

## Acknowledgements

We are grateful for the members of Jain Lab for helpful advice and discussions. We thank the University of Florida, the UF Health Cancer Center, and the Clinical and Translational Science Institute Repository for their support. We also thank Dr. David Ostrov and Dr. Cuong Nguyen for their support with obtaining the patient samples and for their discussions. Finally, we thank Dr. Whitney Stoppel and her lab in the Department of Chemical Engineering and Dr. Chrisopher Dervinis in the Forest Genomics Group at the University of Florida for the use of the lyophilizers. The following reagents were obtained through BEI Resources, NIAID, NIH: Genomic RNA from the Middle East Respiratory Syndrome Coronavirus (MERS-CoV), EMC/2012, NR-45843; Quantitative PCR (qPCR) Control RNA from Inactivated SARS Coronavirus, Urbani, NR-52346; Genomic RNA from Human Coronavirus (HCoV), NL63, NR-44105; Genomic RNA from SARS-Related Coronavirus 2, Isolate Hong Kong/VM20001061/2020, NR-52388. The following reagent was deposited by the Centers for Disease Control and Prevention and obtained through BEI Resources, NIAID, NIH: Genomic RNA from SARS-Related Coronavirus 2, Isolate USA-WA1/ 2020, NR-52285; Quantitative PCR (qPCR) Control RNA from Heat-Inactivated SARS-Related Coronavirus 2, Isolate USA-WA1/2020, NR 52347; SARS-Related Coronavirus 2, Isolate USA-WA1/2020, Heat Inactivated, NR-52286. The following reagent was obtained from Dr. Maria R. Capobianchi through BEI Resources, NIAID, NIH: Genomic RNA from SARS-Related Coronavirus 2, Isolate Italy-INMI1, NR-52498.

