## Supplementary Information for "Engineered CRISPR/Cas12a Enables Rapid SARS-CoV-2 Detection"

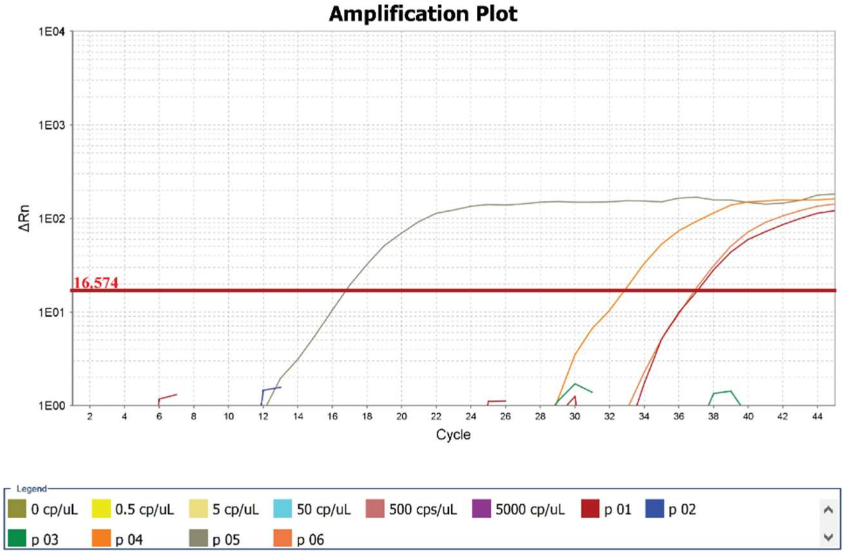

N1 Gene

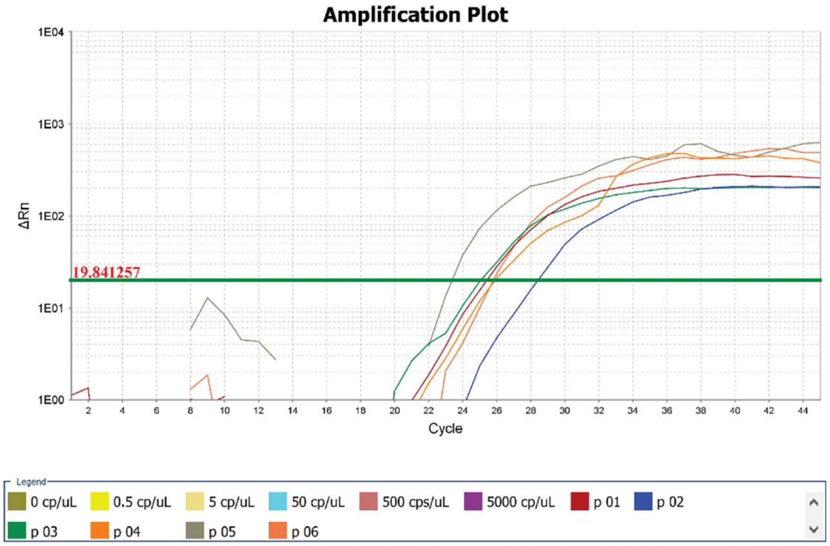

RNase P Gene

**Fig. S1. Representation of clinical validation of SARS-CoV-2 detection in 62 patient samples using RT-qPCR.** Obtained patient samples were re-tested with RT-qPCR following CDC-recommended protocol to determine the Ct values.

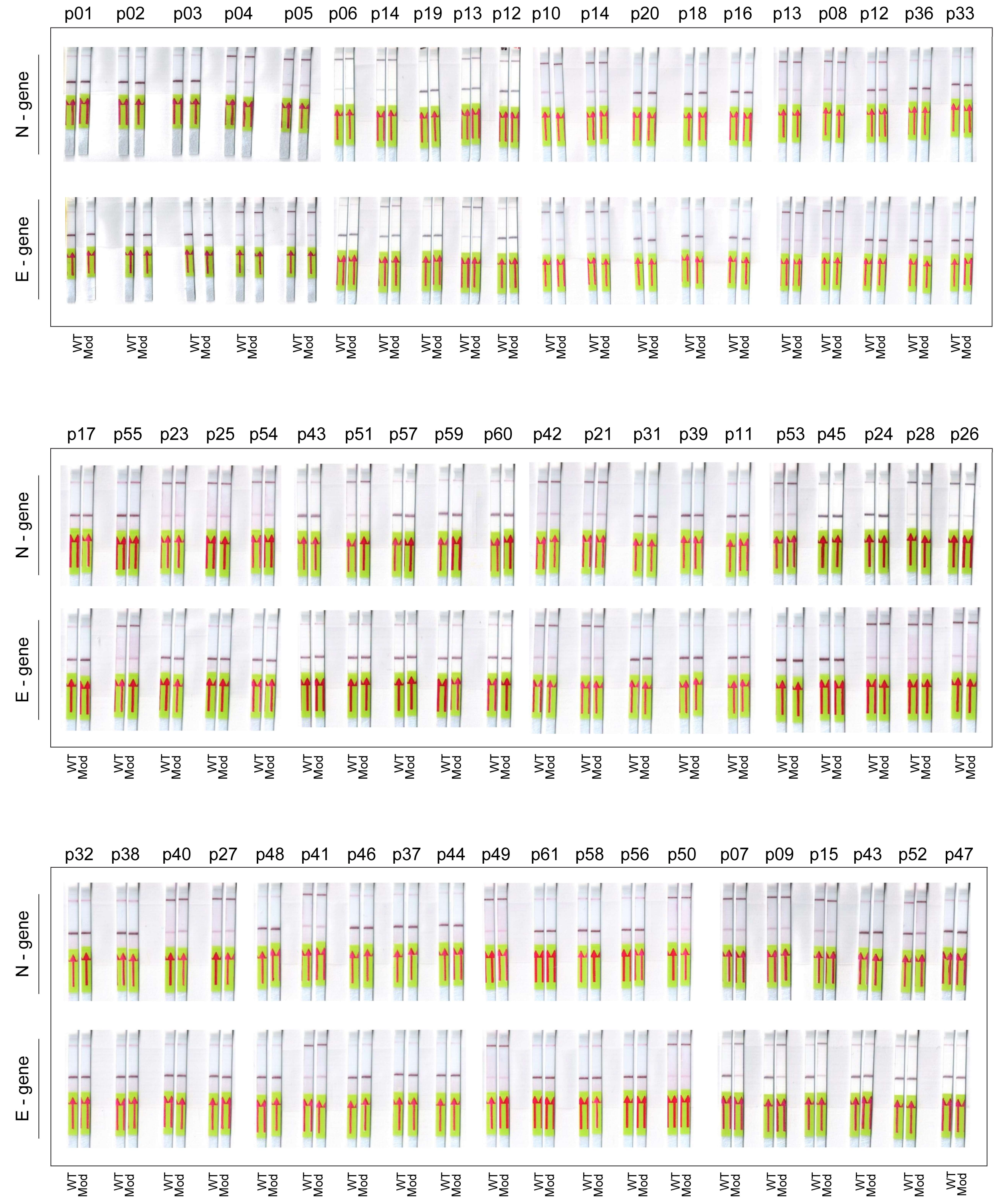

**Fig. S2. Clinical validation of ENHANCE for the detection of SARS-CoV-2 in 62 patient samples using lateral flow assay.** The patient samples were not in numerical order due to blind testing.

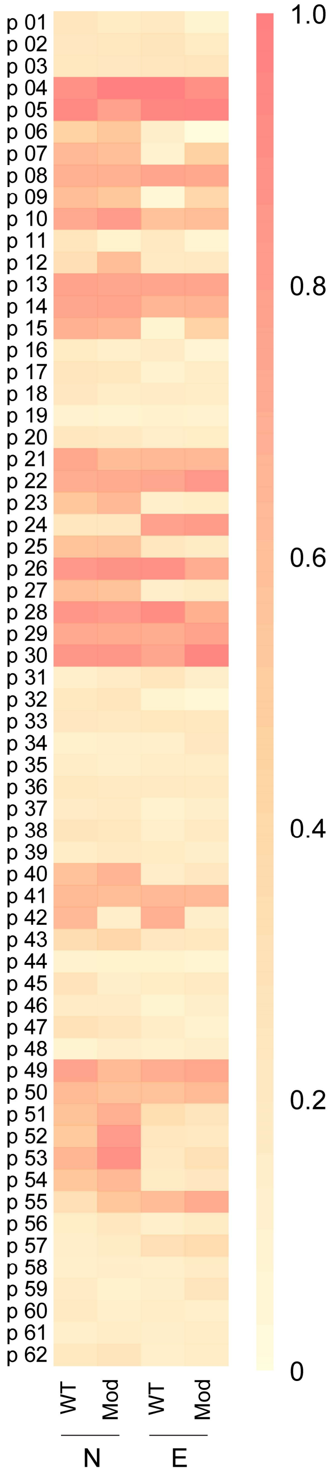

**Fig. S3. Quantitative analysis of lateral flow assay using ENHANCE on 62 patient samples.** The heat map shows ImageJ quantification of band intensity ratio of positive line (top band) to highest signal obtained in all 31 positive patient samples.

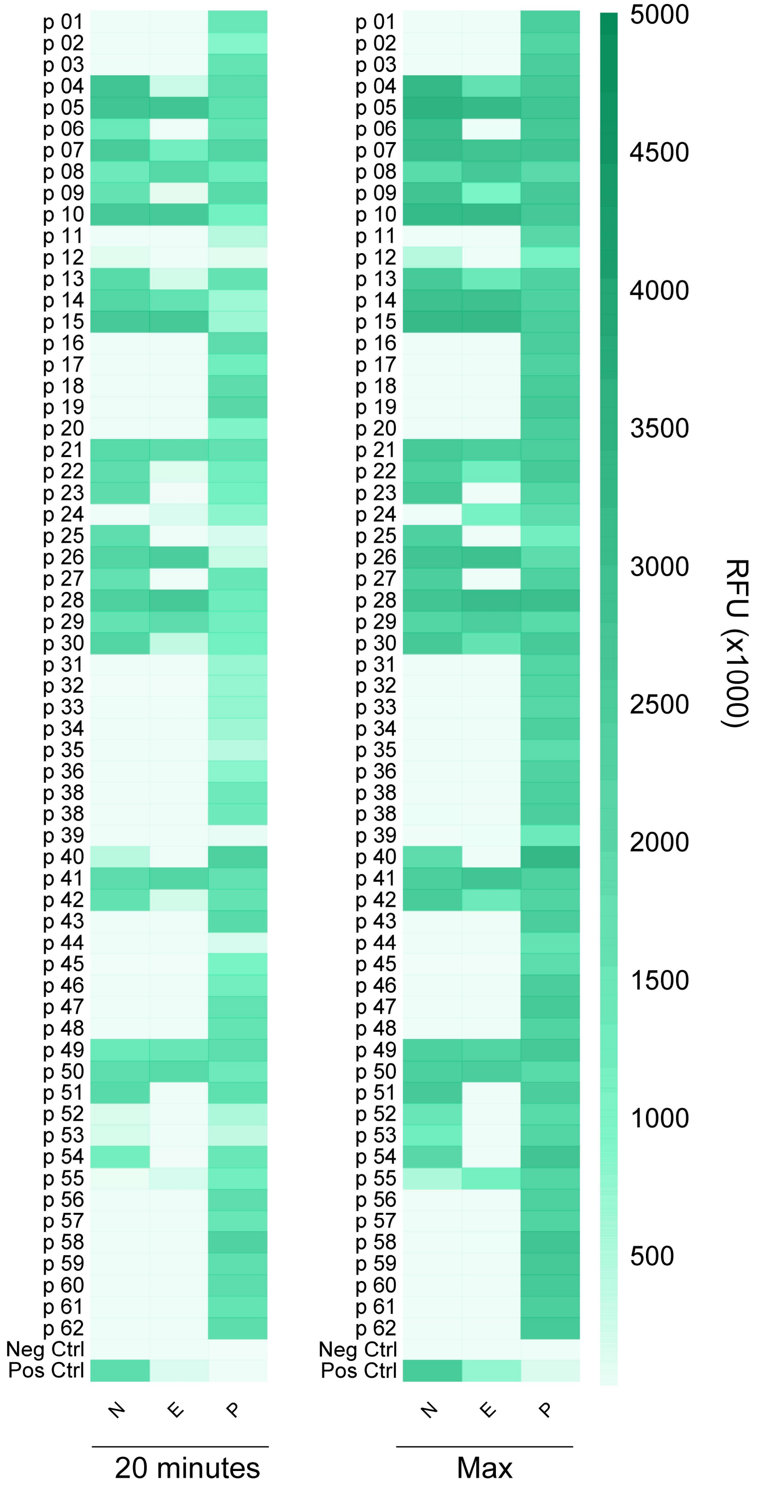

**Fig. S4. Clinical validation of ENHANCEv2 for the detection of SARS-CoV-2 in 62 patient samples using fluorescence-based reporter assay.** The heat map shows fluorescence intensities taken at t = 20 minutes (left) and maximum fluorescence intensities within an hour. The heat map is supplemental to fig. 4e in the main text whose fluorescence intensities were taken at t = 2.5 minutes.

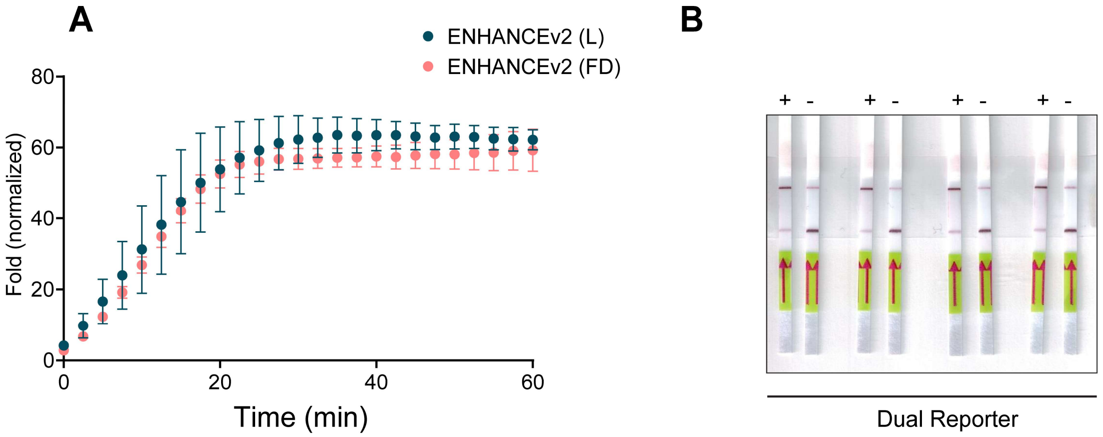

**Fig. S5. Comparison between liquid version and lyophilized version of ENHANCEv2. (A)** Fold change in fluorescence intensity normalized to corresponding NTC reactions targeting N2gene between liquid ENHANCEv2 (L) and freeze-dried ENHANCEv2 (FD) in one hour. (**B**) Representation of lateral flow paper strips showing compatibility of the dual reporter. Sample reactions from the fluorescence-based reporter assay using dual reporter in ENHANCEv2 was diluted down to a final concentration of 125 nM reporter followed by the lateral flow assay.

**Table S1. Comparison of CRISPR-based detection methods**

| | DETECTR | SHERLOCK | StopCOVID.v2 | RT-AIOD | ENHANCE |
|---|---|---|---|---|---|
| **Target genes** | N, E | N, ORF1ab | N | N | N, E |
| **Enzyme** | Cas12a | Cas13a | Cas12b | Cas12a | Cas12a |
| **LoD** copies/μL<br><br>**(Total copies)** | 10-100 copies/μL<br><br>(20 copies input) | 0.9-6.75 copies/μL<br><br>(7-54 copies input) | 0.033 copies/μL<br><br>(100 copies per sample) | 4.6 copies/μL<br><br>(5 copies input) | 0.2-7.9 copies/μL<br><br>(1-40 copies input ) |
| **Time to detect patient samples**<br><br>**(RNA to detection)** | 30-40 min | 60 min | 45-80 min | 40 min | 33 min |
| **Positive Prediction Rate** | 95% | 96.30% | 98.40% | 100% | 96.7% |
| **Negative Prediction rate** | 100% | 100% | 93.40% | 100% | 96.7% |

**Table S2. Sequences used in the study**

| Name | Sequence | Source |
|---|---|---|
| **crRNAs** | | |
| crN1-WT | UAAUUUCUACUAAGUGUAGAUGUGGACCC UAGAUUCAACU | This study |
| crN1-Mod | UAAUUUCUACUAAGUGUAGAUGUGGACCC UAGAUUCAACUTATTATT | This study |
| crN2-WT | UAAUUUCUACUAAGUGUAGAUCCCCCAGC GCUUCAGCGUUC | J.P. Broughton et al. |
| crN2-Mod | UAAUUUCUACUAAGUGUAGAUCCCCCAGC GCUUCAGCGUUCTATTATT | This study |
| crE1-WT | UAAUUUCUACUAAGUGUAGAUGUGGUAUU CUUGCUAGUUAC | J.P. Broughton et al. |
| crE1-Mod | UAAUUUCUACUAAGUGUAGAUGUGGUAUU CUUGCUAGUUACTATTATT | This study |
| crE2-WT | UAAUUUCUACUAAGUGUAGAUUUGCUUUC GUGGUAUUCUUG | J.P. Broughton et al. |
| crE2-Mod | UAAUUUCUACUAAGUGUAGAUUUGCUUUC GUGGUAUUCUUGTATTATT | This study |
| crR1-WT | UAAUUUCUACUAAGUGUAGAUUCAAGCUG UCACGGCCAAUG | This study |
| crR1-Mod | UAAUUUCUACUAAGUGUAGAUUCAAGCUG UCACGGCCAAUGTATTATT | This study |
| crR2-WT | UAAUUUCUACUAAGUGUAGAUACAUUUGU CAAGCUGUCACG | This study |
| crR2-Mod | UAAUUUCUACUAAGUGUAGAUACAUUUGU CAAGCUGUCACGTATTATT | This study |
| crP-WT | UAAUUUCUACUAAGUGUAGAUAAUUACUU GGGUGUGACCCU | K.A Curtis et al. |
| crP-Mod | UAAUUUCUACUAAGUGUAGAUAAUUACUU GGGUGUGACCCUTATTATT | This study |
| **LAMP Primers** | | |
| F3-N1 | TCATGACGTTCGTGTTGT | This study |

| | | |
|---|---|---|
| B3-N1 | TTGAGTGAGAGCGGTGAA | This study |
| FIP-N1 | TAATGCGGGGTGCATTTCGAGATTTCATCT AAACGAACAAAC | This study |
| BIP-N1 | TAACCAGAATGGAGAACGCAAGTATTATTG GGTAAACCTTGG | This study |
| LF-N1 | CTGATTTTGGGGTCCATTA | This study |
| LB-N1 | GTGGGGCGCGATCAAAACAAC | This study |
| F3-N2 | AACACAAGCTTTCGGCAG | J.P. Broughton et al. |
| B3-N2 | GAAATTTGGATCTTTGTCATCC | J.P. Broughton et al. |
| FIP-N2 | TGCGGCCAATGTTTGTAATCAGCCAAGGAA ATTTTGGGGAC | J.P. Broughton et al. |
| BIP-N2 | CGCATTGGCATGGAAGTCACTTTGATGGC ACCTGTGTAG | J.P. Broughton et al. |
| LF-N2 | TTCCTTGTCTGATTAGTTC | J.P. Broughton et al. |
| LB-N2 | ACCTTCGGGAACGTGGTT | J.P. Broughton et al. |
| F3-E1&E2 | CCGACGACGACTACTAGC | J.P. Broughton et al. |
| B3-E1&E2 | AGAGTAAACGTAAAAAGAAGGTT | J.P. Broughton et al. |
| FIP-E1&E2 | CTAGCCATCCTTACTGCGCTACTCACGTTA ACAATATTGCA | J.P. Broughton et al. |
| BIP-E1&E2 | ACCTGTCTCTTCCGAAACGAATTTGTAAGC ACAAGCTGATG | J.P. Broughton et al. |
| LF-E1&E2 | TCGATTGTGTGCGTACTGC | J.P. Broughton et al. |
| LB-E1&E2 | TGAGTACATAAGTTCGTAC | J.P. Broughton et al. |
| F3-R1&R2 | TCAAGTATTGAGTGAAATGGTC | This study |
| B3-R1&R2 | TCTATAGAGACACTCATAAAGTCT | This study |
| FIP-R1&R2 | AAACACTATTAGCATAAGCACGGTTCACTA TATGTTAAACCA | This study |
| BIP-R1&R2 | TGCACTTTTATCTACTGATGGTGTTGTAAAT TGCGGACAT | This study |
| LF-R1&R2 | GTTGTGGCATCTCCTGATGA | This study |
| LB-R1&R2 | GTAACAAAATTGCCGATAAG | This study |

| | | |
|---|---|---|
| F3-RNaseP | TTGATGAGCTGGAGCCA | K.A Curtis et al. |
| B3-RNaseP | CACCCTCAATGCAGAGTC | K.A Curtis et al. |
| FIP-RNaseP | GTGTGACCCTGAAGACTCGGTTTTAGCCA CTGACTCGGATC | K.A Curtis et al. |
| BIP-RNaseP | CCTCCGTGATATGGCTCTTCGTTTTTTTCTT ACATGGCTCTGGTC | K.A Curtis et al. |
| LF-RNaseP | ATGTGGATGGCTGAGTTGTT | K.A Curtis et al. |
| LB-RNaseP | CATGCTGAGTACTGGACCTC | K.A Curtis et al. |
| **Reporter** | | |
| Fluorescence-based reporter | /56-FAM/TTATT/3IABkFQ/ | J.S. Chen et al |
| Lateral flow reporter | /56-FAM/TTATTATT/3Bio/ | J.P. Broughton et al. |
| Dual reporter | /56-FAM/TTATTA/iBiodT/3IABkFQ/ | This study |
| **LbCas12a amino acid sequences** | | |
| LbCas12a | MSKLEKFTNCYSLSKTLRFKAIPVGKTQENID NKRLLVEDEKRAEDYKGVKKLLDRYYLSFIN DVLHSIKLKNLNNYISLFRKKTRTEKENKELE NLEINLRKEIAKAFKGNEGYKSLFKKDIIETILP EFLDDKDEIALVNSFNGFTTAFTGFFDNREN MFSEEAKSTSIAFRCINENLTRYISNMDIFEKV DAIFDKHEVQEIKEKILNSDYDVEDFFEGEFF NFVLTQEGIDVYNAIIGGFVTESGEKIKGLNE YINLYNQKTKQKLPKFKPLYKQVLSDRESLSF YGEGYTSDEEVLEVFRNTLNKNSEIFSSIKKL EKLFKNFDEYSSAGIFVKNGPAISTISKDIFGE WNVIRDKWNAEYDDIHLKKKAVVTEKYEDDR RKSFKKIGSFSLEQLQEYADADLSVVEKLKEII IQKVDEIYKVYGSSEKLFDADFVLEKSLKKND AVVAIMKDLLDSVKSFENYIKAFFGEGKETNR DESFYGDFVLAYDILLKVDHIYDAIRNYVTQK PYSKDKFKLYFQNPQFMGGWDKDKETDYR ATILRYGSKYYLAIMDKKYAKCLQKIDKDDVN GNYEKINYKLLPGPNKMLPKVFFSKKWMAYY NPSEDIQKIYKNGTFKKGDMFNLNDCHKLIDF FKDSISRYPKWSNAYDFNFSETEKYKDIAGF YREVEEQGYKVSFESASKKEVDKLVEEGKL YMFQIYNKDFSDKSHGTPNLHTMYFKLLFDE NNHGQIRLSGGAELFMRRASLKKEELVVHPA NSPIANKNPDNPKKTTTLSYDVYKDKRFSED | M.A. Moreno-Mateos et al |

| | | |
|---|---|---|
| | QYELHIPIAINKCPKNIFKINTEVRVLLKHDDN<br>PYVIGIDRGERNLLYIVVVDGKGNIVEQYSLN<br>EIINNFNGIRIKTDYHSLLDKKEKERFEARQN<br>WTSIENIKELKAGYISQVVHKICELVEKYDAVI<br>ALEDLNSGFKNSRVKVEKQVYQKFEKMLIDK<br>LNYMVDKKSNPCATGGALKGYQITNKFESFK<br>SMSTQNGFIFYIPAWLTSKIDPSTGFVNLLKT<br>KYTSIADSKKFISSFDRIMYVPEEDLFEFALDY<br>KNFSRTDADYIKKWKLYSYGNRIRIFRNPKKN<br>NVFDWEEVCLTSAYKELFNKYGINYQQGDIR<br>ALLCEQSDKAFYSSFMALMSLMLQMRNSITG<br>RTDVDFLISPVKNSDGIFYDSRNYEAQENAIL<br>PKNADANGAYNIARKVLWAIGQFKKAEDEKL<br>DKVKIAISNKEWLEYAQTSVKH | |
| LbCas12a$^{D156R}$ | MSKLEKFTNCYSLSKTLRFKAIPVGKTQENID<br>NKRLLVEDEKRAEDYKGVKKLLDRYYLSFIN<br>DVLHSIKLKNLNNYISLFRKKTRTEKENKELE<br>NLEINLRKEIAKAFKGNEGYKSLFKKDIIETILP<br>EFLDDKDEIALVNSFNGFTTAFTGFF**R**NREN<br>MFSEEAKSTSIAFRCINENLTRYISNMDIFEKV<br>DAIFDKHEVQEIKEKILNSDYDVEDFFEGEFF<br>NFVLTQEGIDVYNAIIGGFVTESGEKIKGLNE<br>YINLYNQKTKQKLPKFKPLYKQVLSDRESLSF<br>YGEGYTSDEEVLEVFRNTLNKNSEIFSSIKKL<br>EKLFKNFDEYSSAGIFVKNGPAISTISKDIFGE<br>WNVIRDKWNAEYDDIHLKKKAVVTEKYEDDR<br>RKSFKKIGSFSLEQLQEYADADLSVVEKLKEII<br>IQKVDEIYKVYGSSEKLFDADFVLEKSLKKND<br>AVVAIMKDLLDSVKSFENYIKAFFGEGKETNR<br>DESFYGDFVLAYDILLKVDHIYDAIRNYVTQK<br>PYSKDKFKLYFQNPQFMGGWDKDKETDYR<br>ATILRYGSKYYLAIMDKKYAKCLQKIDKDDVN<br>GNYEKINYKLLPGPNKMLPKVFFSKKWMAYY<br>NPSEDIQKIYKNGTFKKGDMFNLNDCHKLIDF<br>FKDSISRYPKWSNAYDFNFSETEKYKDIAGF<br>YREVEEQGYKVSFESASKKEVDKLVEEGKL<br>YMFQIYNKDFSDKSHGTPNLHTMYFKLLFDE<br>NNHGQIRLSGGAELFMRRASLKKEELVVHPA<br>NSPIANKNPDNPKKTTTLSYDVYKDKRFSED<br>QYELHIPIAINKCPKNIFKINTEVRVLLKHDDN<br>PYVIGIDRGERNLLYIVVVDGKGNIVEQYSLN<br>EIINNFNGIRIKTDYHSLLDKKEKERFEARQN<br>WTSIENIKELKAGYISQVVHKICELVEKYDAVI<br>ALEDLNSGFKNSRVKVEKQVYQKFEKMLIDK<br>LNYMVDKKSNPCATGGALKGYQITNKFESFK<br>SMSTQNGFIFYIPAWLTSKIDPSTGFVNLLKT<br>KYTSIADSKKFISSFDRIMYVPEEDLFEFALDY<br>KNFSRTDADYIKKWKLYSYGNRIRIFRNPKKN<br>NVFDWEEVCLTSAYKELFNKYGINYQQGDIR | This study |

| | |
|---|---|
| | ALLCEQSDKAFYSSFMALMSLMLQMRNSITGRTDVDFLISPVKNSDGIFYDSRNYEAQENAILPKNADANGAYNIARKVLWAIGQFKKAEDEKLDKVKIAISNKEWLEYAQTSVKH |

**Table S3. Interpretation of results**

**Fluorescence-based reporter detection assay**

**Table S3.1 Assay Controls**

| | Positive Control | Negative Control | Interpretation |
|---|---|---|---|
| **SARS-CoV-2 N2 Gene ($N2_{t=20}/N2_{t=0}$)** | ≥ 5 | < 5 | Valid for N2 gene. |
| | < 5 | < 5 | Invalid for positive control. Indicates QC failure. |
| | ≥ 5 | ≥ 5 | Invalid for negative control. Indicates N2 gene contamination |
| **SARS-CoV-2 E2 Gene ($E2_{t=20}/E2_{t=0}$)** | ≥ 5 | < 5 | Valid for E2 gene. |
| | < 5 | < 5 | Invalid for positive control. Indicates QC failure. |
| | ≥ 5 | ≥ 5 | Invalid for negative control. Indicates E2 gene contamination |
| **Human RNASE-P Gene ($RNASE\text{-}P_{t=20}$ / $RNASE\text{-}P_{t=0}$)** | N/A | < 5 | Valid for RNASE-P. |
| | | ≥ 5 | Invalid. Indicates RNASE-P gene contamination. |

**Table S3.2 Fluorescence Fold-Change Interpretation**

| **SARS-CoV-2 N2 Gene ($N2_{t=20}$ /$NTC_{t=20}$)** | **SARS-CoV-2 E2 Gene ($E2_{t=20}$ /$NTC_{t=20}$)** | **Human RNASE-P Gene ($RNASE\text{-}P_{t=20}$ /$NTC_{t=20}$)** | **Interpretation** |
|---|---|---|---|
| ≥ 5 | ≥ 5 | N/A | Positive for SARS-CoV-2 |
| ≥ 5 | < 5 | N/A | |
| < 5 | ≥ 5 | N/A | |
| < 5 | < 5 | ≥ 5 | Negative for SARS-CoV-2 |

**Lateral Flow assay**

**Table S3.3 Assay Controls**

| | Positive Control | Negative Control | Interpretation |
|---|---|---|---|
| **SARS-CoV-2 N2 Gene Visual readout** | +<br><br>-<br><br>+ | -<br><br>+<br><br>+ | Valid for N2 gene.<br><br>Invalid for positive control. Indicates QC failure.<br><br>Invalid for negative control. Indicates N2 gene contamination |
| **SARS-CoV-2 E2 Gene Visual readout** | +<br><br>-<br><br>+ | -<br><br>+<br><br>+ | Valid for E2 gene.<br><br>Invalid for positive control. Indicates QC failure.<br><br>Invalid for negative control. Indicates E2 gene contamination |
| **Human RNASE-P Gene Visual readout** | N/A | -<br><br>+ | Valid for RNASE-P.<br><br>Invalid. Indicates RNASE-P gene contamination. |

**Table S3.4 Lateral Flow Sample Interpretation**

| SARS-CoV-2 N2 Gene Visual Readout | SARS-CoV-2 E2 Gene Visual Readout | Human RNASE-P Gene Visual Readout | Interpretation |
|---|---|---|---|
| + | + | N/A | Positive for SARS-CoV-2 |
| + | - | N/A | |
| - | + | N/A | |
| - | - | + | Negative for SARS-CoV-2 |
